# Changes in social behavior over time in the COVID-19 pandemic

**DOI:** 10.1101/2020.08.27.20183376

**Authors:** Megan M. Sheehan, Elizabeth R. Pfoh, Sidra Speaker, Michael B. Rothberg

## Abstract

Public health recommendations aimed at limiting spread of SARS-CoV-2 have encouraged social distancing and masks as economies across the United States re-open. Understanding adherence to these guidelines will inform further efforts to reduce transmission. In this repeated cross-sectional survey study, we describe changes in social behavior in Ohio during periods of declining and rising cases. While essential activities remained consistent over time, more individuals attended gatherings of 10 or more people as cases rose, particularly in the 18-29 age group. A majority of individuals wore masks. It appears necessary to continue limiting gatherings and encourage mask-wearing, particularly among younger groups.

## Introduction

In March 2020, Ohio and other states closed schools, workplaces, and gathering spots to limit transmission of SARS-CoV-2. Gatherings of more than 10 people were largely prohibited, and the adoption of social distancing and cloth face masks was encouraged to prevent rapid spread of the virus^1^. As states flattened the curve of new transmissions, they began to reopen. Unfortunately, as restrictions lifted, cases have increased^2^. Understanding to what extent individuals adhered to public health recommendations during reopening can help inform current efforts to shape public behavior and reduce transmission. In this study, we describe social behavior in Ohio early in the re-opening phase and then afterward as cases rose.

## Methods

This repeated cross-sectional study surveyed adults seen at a large integrated health system in Ohio in the past year and active on MyChart. We excluded patients who tested positive for SARS-COV2 prior to the survey. Patients were sent a 14-question survey via MyChart about social distancing and hygiene behaviors in the past 7 days. The survey was developed based on expert opinion and prior surveys^3,4^. It was pilot tested for clarity and sent out in seven waves from May 19 to July 24. After an initial lock-down period, Ohio began to reopen on April 30; daily cases bottomed on June 15 then rose substantially; mask-wearing was mandated on July 23.

Survey responses before and after the June 15 nadir were compared using Student’s t-tests for numeric variables and Pearson chi-square or Fisher’s Exact tests for categorical variables. Secondarily, we compared respondents who stated they “always wore masks” to respondents who wore masks less often to identify if there were other differences in public health behaviors. All statistical tests were two-tailed with a significance threshold of 0.05. Analyses were conducted using R v.4.02. This work was approved by Cleveland Clinic Institutional Review Board.

## Results

A total of 654 individuals responded (7% response rate) and were similar in age and sex before and after June 15. A majority of respondents wore a mask outside the home, 53% before and 64% after (p = 0.008). More respondents attended a gathering of 10 or more people after June 15 (19% versus 11%, p = 0.01) (Table 1). Mask wearing was less common among those who attended a large gathering than those who did not (34% versus 66%).

**Table 1.** Survey responses before and after the nadir of cases.* *Survey responses were grouped into before the nadir of COVID-19 cases in Ohio (June 15, 2020) and after. Besides the demographic and health status questions, the survey asked respondents to use a 7-day look-back period for their activities. Bolding indicates statistical significance ≤0.02.

| *Survey Question* | *Pre-nadir*<br>*n=214*<br>*N (%)* | *Post-nadir*<br>*n=440*<br>*N (%)* | *p-value* |
| --- | --- | --- | --- |
| Close contact with someone who has COVID* | 1 (0.5) | 5 (1) | 0.67 |
| *Demographics and health status* |  |  |  |
| Age | 54 (37, 66) | 56 (36, 70) | 0.53 |
| Gender - Female | 139 (66) | 304 (70) | 0.33 |
| Number of people living in the home | 2 (2, 4) | 2 (2, 3) | 0.41 |
| Someone in the home had URI symptoms in past week | 7 (3) | 20 (5) | 0.54 |
| Someone in the home provides patient care | 24 (11) | 38 (9) | 0.35 |
| Been placed in isolation or quarantine | 9 (4) | 8 (2) | 0.11 |
| Lung disease (e.g. asthma, chronic obstructive pulmonary disease) | 30 (14) | 61 (14) | 1 |
| Immune suppression | 20 (9) | 32 (7) | 0.44 |
| High blood pressure | 65 (30) | 127 (29) | 0.76 |
| *Social activities* |  |  |  |
| Gone to a friend, neighbor, or relative's residence | 108 (51) | 211 (48) | 0.64 |
| **Attended a large gathering (10+ people)** | **23 (11)** | **84 (19)** | **0.01** |
| Gone out to a bar, club, or other place where people gather | 26 (12) | 66 (15) | 0.38 |
| *Activities of daily living* |  |  |  |
| Gone to the grocery store or pharmacy | 167 (79) | 342 (78) | 0.88 |
| Sought care from a hospital or health care facility | 35 (16) | 95 (22) | 0.13 |
| Remained in your residence at all times, except for essential activities or exercise | 122 (57) | 243 (55) | 0.72 |
| Shared items like towels or utensils with other people | 55 (26) | 84 (19) | 0.06 |
| Had close contact (within 6 feet) with people who live with you | 181 (85) | 359 (83) | 0.50 |
| Had close contact (within 6 feet) with people who do not live with you | 125 (58) | 230 (53) | 0.20 |
| Gone outside to walk, hike, or exercise | 182 (86) | 351 (81) | 0.16 |
| Shared a household with someone who has been interacting with 10+ people/day. | 35 (16) | 103 (23) | 0.05 |
| *Travel and work* |  |  |  |
| Used shared public transit (e.g. train, bus or subway) | 1 (0.5) | 7 (2) | 0.28 |
| Used a ride-share service (e.g. Uber or Lyft) or Taxi. | 2 (1) | 8 (2) | 0.51 |
| Travelled by airplane | 0 (0) | 2 (0.5) | 1 |
| Worked or studied outside the home | 55 (26) | 118 (27) | 0.83 |
| Been exposed to 10 or more people per day through my work. | 39 (18) | 85 (19) | 0.82 |
| *Cleaning* |  |  |  |
| ALWAYS washed my hands or used hand sanitizer after possible exposures | 130 (62) | 293 (67) | 0.21 |
| ALWAYS washed my hands for at least 20 seconds | 157 (74) | 304 (69) | 0.28 |
| ALWAYS cleaned and disinfected shared surfaces daily | 92 (43) | 183 (42) | 0.87 |
| **ALWAYS cleaned or disinfected mail or groceries.** | **53 (25)** | **74 (17)** | **0.02** |
| *Risk avoidance* |  |  |  |
| ALWAYS worn gloves outside of home. | 15 (7) | 20 (5) | 0.21 |
| ALWAYS avoided leaving my home at all | 21 (10) | 36 (8) | 0.58 |
| ALWAYS avoided others in public spaces | 76 (36) | 177 (41) | 0.29 |
| ALWAYS avoided touching my face while in public spaces. | 92 (44) | 222 (51) | 0.08 |
| **ALWAYS wore a mask when leaving home** | **113 (53)** | **281 (64)** | **0.008** |
| Which type of mask did you wear? |  |  |  |
| **Cloth** | **141 (66)** | **329 (75)** | **0.02** |
| Surgical | 82 (38) | 135 (31) | 0.06 |
| N95 | 22 (10) | 41 (9) | 0.80 |
| Other | 10 (5) | 15 (3) | 0.57 |

Changes in social behavior differed by age group. Public gatherings increased among 18-29-year-olds and 30-49-year-olds (by 25%, p=0.02 and 17%, p=0.05, respectively), but not among older patients. Mask usage increased among 50-64-year-olds (40% to 58%, p=0.03).

Respondents over 65 had the highest mask usage during both time periods. Activities such as grocery shopping, seeking healthcare, and hand-washing did not change over time in any group (p>0.07) (Table 2).

**Table 2.**
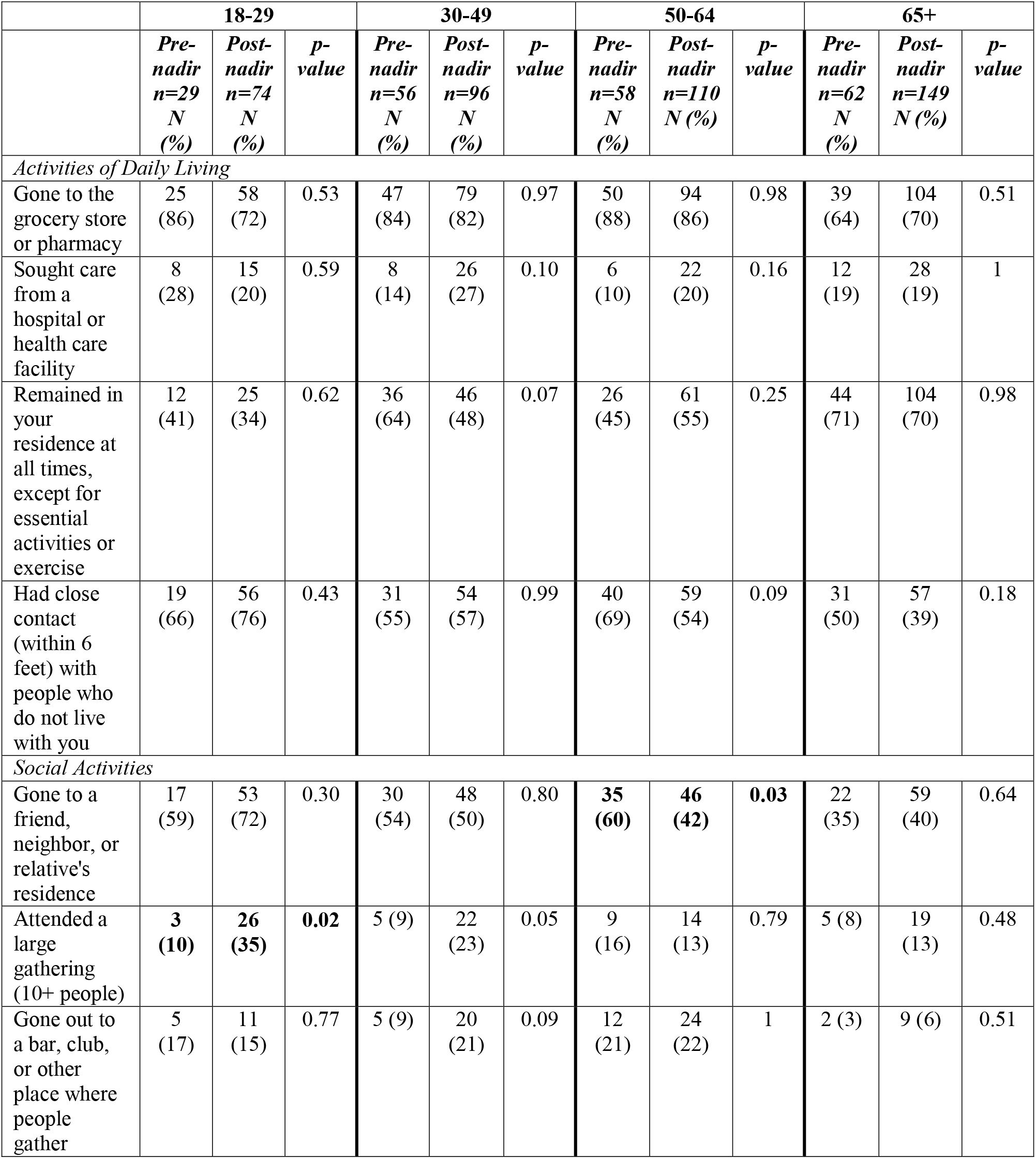
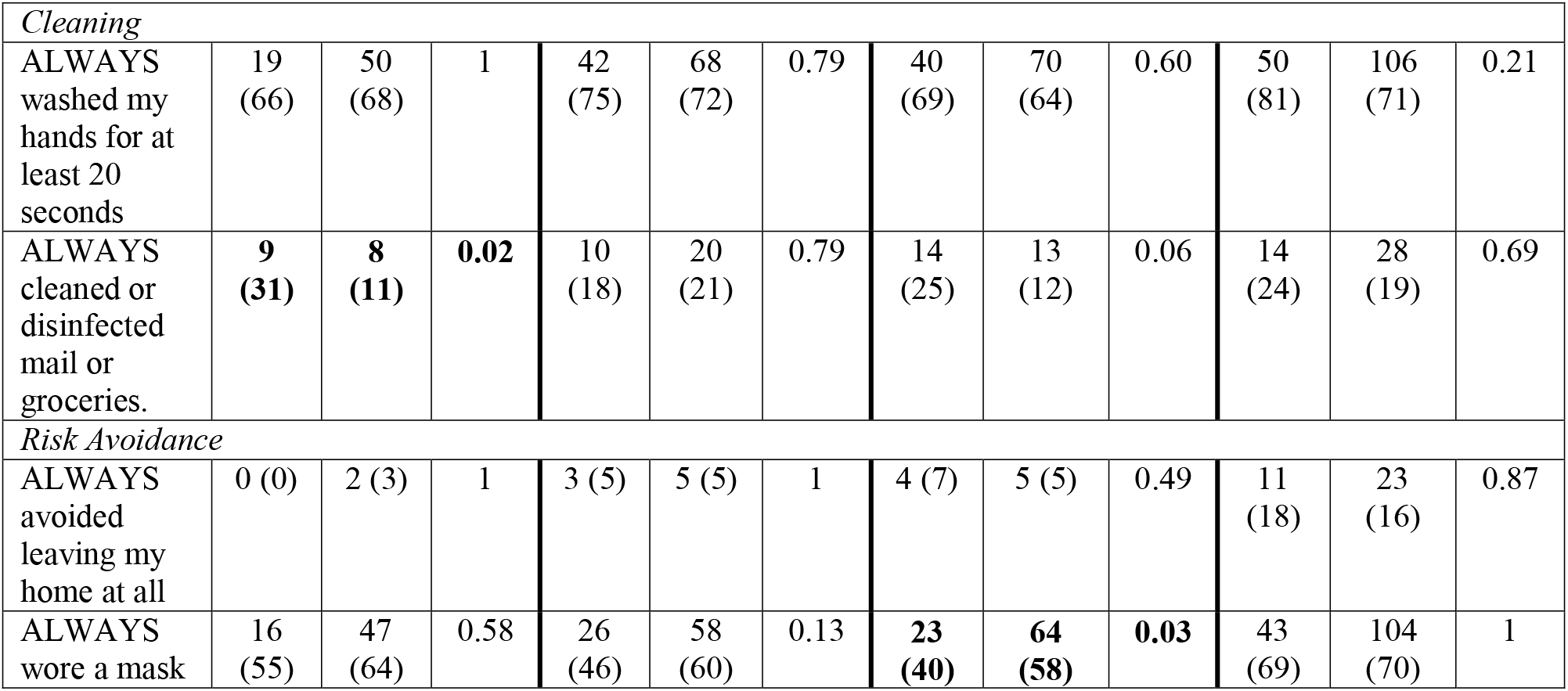
Activities of daily living, social activities, and risk avoidance before and after the nadir of cases by age group.* *Survey responses were grouped into before the nadir of COVID-19 cases in Ohio (June 15, 2020) and after. Besides the demographic and health status questions, the survey asked respondents to use a 7-day look-back period for their activities. Bolding indicates statistical significance <0.05.

Respondents who always wore masks practiced other protective behaviors. Those who always masked were more often female (75% versus 59%), washed hands more frequently (78% versus 46%) and avoided others in public spaces (51% versus 22%). Respondents who did not always mask outside their home were more likely to visit a friend or neighbor’s residence (58% versus 43%) or a bar or club (24% versus 8%), and have close contact with people that they did not live with (70% versus 45%).

## Discussion

In this repeated cross-sectional survey study, we found an increasing percentage of individuals attended large gatherings as cases were rising in Ohio, compared to a period of declining cases. Increased socialization was most apparent among 18-29-year-olds, possibly illustrating ‘caution fatigue’ amongst those at lowest risk of dying from COVID-19^5^. This behavior has been reported anecdotally in the lay press, but has not been previously documented. While the majority of respondents, especially those over 65 years, used masks, individuals who attended gatherings did so less consistently. Essential activities and sanitization practices remained largely consistent across time and age groups.

A recent review concluded that face masks do not adversely affect hand hygiene^6^. Our study supports this finding, as respondents who wore masks more often followed all public health recommendations.

Study limitations include sampling from a population that has accessed the healthcare system in the past year and may be more aware of public health messaging.

As the national conversation focuses on safe economic revival, it appears important to limit gatherings of more than 10 people and encourage mask-wearing. Messaging should target younger patients, who are also least likely to wear masks.

## Data Availability

The data that support the findings of this study are available from the corresponding author, upon reasonable request.

## Acknowledgment

We would like to thank Gina Rupp, RN, MSN and Oleg Lisheba for their assistance in data collection.

The authors have no conflicts of interest to disclose.

## References

1. Schuchat A. Public Health Response to the Initiation and Spread of Pandemic COVID-19 in the United States, February 24-April 21, 2020. MMWR Morb Mortal Wkly Rep. 2020;69. doi:10.15585/mmwr.mm6918e2

2. CDC. Coronavirus Disease 2019 (COVID-19) in the U.S. Centers for Disease Control and Prevention. Published July 12, 2020. Accessed July 14, 2020. https://www.cdc.gov/coronavirus/2019-ncov/cases-updates/cases-in-us.html

3. Hamilton CM, Strader LC, Pratt JG, et al. The PhenX Toolkit: get the most from your measures. Am J Epidemiol. 2011;174(3):253–260. doi:10.1093/aje/kwr193

4. Rubin GJ, Amlôt R, Page L, Wessely S. Public perceptions, anxiety, and behaviour change in relation to the swine flu outbreak: cross sectional telephone survey. The BMJ. 2009;339. doi:10.1136/bmj.b2651

5. Verity R, Okell LC, Dorigatti I, et al. Estimates of the severity of coronavirus disease 2019: a model-based analysis. Lancet Infect Dis. 2020;20(6):669–677. doi:10.1016/S1473-3099(20)30243-7

6. Mantzari E, Rubin GJ, Marteau TM. Is risk compensation threatening public health in the covid-19 pandemic? BMJ. 2020;370. doi:10.1136/bmj.m2913

